# Did the National lockdown lock COVID-19 down in India, and reduce pressure on health infrastructure?

**DOI:** 10.1101/2020.05.27.20115329

**Authors:** Zakir Husain, Arup Kumar Das, Saswata Ghosh

## Abstract

**Background & objectives:** The spread of COVID-19 in India has posed a major challenge for policy makers. Policy response in form of imposition of a prolonged national lockdown has imposed substantial costs on the entire population. But the extent to which it has contained the spread of the epidemic needs to be assessed.

**Methods:** We use an Interrupted Time Series model to assess the success of lockdowns in containing COVID-19. In the second step, we use four variants of the SIR models to develop a counterfactual—what would have happened without the lockdown. These results are compared with actual data. The analysis is undertaken for India, and Maharashtra, Gujarat, Delhi, and Tamil Nadu.

**Results:** Lockdown has reduced the number of COVID-19 cases by 23.65 to 337.73 lakh in Class I cities and towns, where COVID has mainly spread. It has averted about 0.01 to 0.10 lakh deaths. At the regional level, however, lockdown has averted a health crisis as existing ICU and ventilator facilities for critically ill patients would have been inadequate.

**Interpretation & conclusions:** Overall, the results for three of the four models reveal that lockdown has a modest impact on spread of COVID-19; the health infrastructure at the national level is not over strained, even at the peak. At the regional level, on the other hand, lockdowns may have been justified. However, given that identification of new cases is limited by levels of daily testing that are low even by Asian standards, analysis based upon official data may have limitations and result in flawed decisions.

## 1. Introduction

This study estimates the impact of the lockdown on the Indian economy, specifically those residing in Class I cities, to contain the spread of 2019-nCov—popularly referred to as COVID-19. Specifically, we have estimated the effect of the lockdown in terms of reducing number of COVID-19 cases, deaths, and pressure on health infrastructure in India.

COVID-19 is a group of viruses affecting human beings through zoonotic transmission. COVID-19 was manifested in the Province of Hubei, China in December 2019. By 13^th^ March, the outbreak had spread to 114 countries with more than 118,000 cases and 4,291 deaths, leading WHO to declare it a pandemic^1^. The major reason for concern with COVID-19 is its the global scale of transmission, significant number of deaths, infection and mortality of care providers and healthcare workers (HCWs), and higher risk of death in vulnerable or susceptible groups^2^. In such circumstances, particularly in the absence of a licensed vaccine or effective therapeutics for COVID-19, slowing or breaking the transmission dynamics through quarantining and social distancing has been adopted as a strategy^3^. Quite a few countries, including India (vide MHA Order No. 40–3/2020-D dated 24/3/2020), have also adopted a lockdown of the economy.

Lockdown imposes a substantial economic and humanitarian burden on societies, particularly in developing countries, that persists even after the epidemic dies out. Such a cost is justified on the grounds that it decreases number of cases, and delays peak, and consequently reduces pressure on health infrastructure. In a press statement made on 23^rd^ May, 2020, the Niti Ayyog claimed that without lockdown there would have been 36–70 lakh cases, and 37–71 thousand deaths^4^. There have been several studies attempting to estimate the impact of travel bans, social distancing, containment and lockdown on flattening the transmission curve using exponential growth models and the SIR model^5^, and its variants. Such attempts compare actual cases with projected cases estimated using SIR models, and its variants. The limitation of such exercises is that parametric assumptions are either data driven (so that noises in the data, particularly due to low testing rates, distort parametric assumptions), or are based upon parameters from studies in other countries (which may not reflect the socio-economic realities of India).

This study estimates a SIR model based on actual number of cases reported for India in the John Hopkins and MoHFW websites as counterfactual. Alternative parametric values are used to estimate the trajectory of COVID-19 without lockdown. While two projections uses parametric values estimated from data, we also present another set using data on daily number of contacts before lockdown obtained from an online survey, along with assumptions based on different studies. We also use an Interrupted Time Series model, commonly used to study the impact on interventions on population level data, to study the impact of the first three lockdowns upon actual spread of cases^6^. Estimates are made at the national level, and for four states with highest COVID-19 cases, containing 68 percent of cases as on 17^th^ May 2020—Maharashtra, Gujarat, Delhi, and Tamil Nadu. Our study contributes to literature in three ways. Firstly, it uses mathematical models with varying assumptions to estimate plausible courses of the pandemic. Secondly, it uses the ITS model to assess the impact of lockdowns. Finally, it examines pressure on health infrastructure. Results indicate that the impact of lockdown on mortality varies, depending upon the parametric values. It calls for sophisticated calibration of models based greater understanding of the transmission dynamics of COVID-19.

## 2. Materials and method

### 2.1 Review of India studies

There have been several studies attempting to estimate the effect of policies like social distancing, quarantining, and lockdown on restricting the spread of COVID-19. Such studies have produced different estimates of the efficacy of lockdowns in containing transmission of COVID-19 cases in India. A study using estimates based on exponential and SIR models reports that symptomatic quarantine can achieve meaningful reductions in peak prevalence, and spread out the outbreak over a longer period even with high R_0_^6^. Mandal and Mandal observes that lockdown had reduced number of cases by 78 percent^8^. A study by IIPS reports that strict implementation of the third lockdown will ensure that confirmed cases at the peak will be only three million; if, however, the R_0_ prevailing during the second lockdown continues, this figure will be 20 million^9^. A study for Kochi, Bengaluru, Mumbai, and Pune using the eSIR model^10^ reports that:

> “without enforcing the interventions, the predicted counts are going to exceed the estimated capacity of hospital beds in India (estimated at 0.7 per 1000). … We also note … for example the predicted number of cases by May 15 will be at 161 per 100,000 without any intervention (2.2 million total cases nationwide) and will reduce to1 per 100,000 (13,800 total cases nationwide) with the most severe form of intervention”^11^ (p. 12).

Comparing projected increase in confirmed cases in absence of lockdown, a study based on an exponential growth model reported that the projected number of confirmed cases would have been 86,373 on 23^rd^ April, compared to actual count of 23,039^12^. The estimated number of COVID-19 cases in government hospitals would be about three per hospital, but this figure has been significantly reduced to 0.82 through Government policies^12^. Another study reported that lockdown decreased R_0_ from 1.862 to 1.2; duration of the epidemic increases from 2 to 6 months, but would die out by November. Only 0.23 percent of the population would be affected^9^. A study using Bayesian analysis, however found that a three-week lockdown would be insufficient to prevent resurgence; a more sustained lockdown with periodic relaxation was suggested^13^.

In any impact evaluation exercise, identifying the counterfactual—what would have happened in the absence of the intervention—is the crucial problem. While the SIR model and its variants have been utilized to estimate the counterfactual scenario, a major problem with this approach is that there are no reliable estimates for the parametric values embedded in the model^14^. For instance, current models in India either do not account for the contact distribution, or end up assuming that this distribution can be extrapolated from Western countries, which are markedly different in their social and living arrangements^15^. Exponential models, for instance, provide a poor fit to the growth in confirmed cases, both in the initial period and subsequently.

The impact of the lockdown on reducing COVID-19 cases in India is estimated in this study by comparing between:

a. *Actual progress of COVID in India with imposition of lock down measures*: We have used actual data available from the covid19india.org website (which compiles information from various sources, and matches with MoHFW figures) on day-to-day basis. we have used a linear Interrupted Time Series (ITS) Model with interruptions corresponding to each of the first three lock downs to assess the impact of lockdowns on number of cases.
b. *Estimated progress of COVID in India in absence of lockdown*: This is “missing”. We have, therefore, used the SIR model to estimate the probable progress of COVID in the absence of lock down – possible number of COVID-19 cases without the lock down.

### 2.2 Interrupted time series (ITS) model

Interrupted time series (ITS) model is a tool suitable for evaluating the effectiveness of a large-scale public health interventions at a population level, in the absence or presence of suitable counterfactuals^6^. It has been widely used in public health interventions such as vaccination, evaluation of health impacts of unplanned global financial crisis, effect of smoking ban in public places on hospital admissions for acute coronary events etc.^16,17^. The ITS model can be presented as:

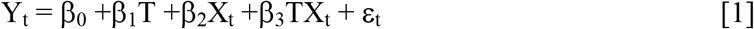

where

Y_t_: the aggregated outcome at time t, in our case the number of new cases of COVID19

T: the time since the start of the study

X_t_: a dummy variable indicating pre-intervention period (coded as 0) or post intervention period (coded as 1)

β_0_ represents the intercept or the initial value of the outcome at T = 0,

β_1_ is the slope or trajectory represents the change in outcome associated with an increase in the time unit (representing the underlying pre-intervention change),

β_2_ represents the level change following the intervention,

β_3_ indicates the slope change following the intervention (using the interaction between time and intervention), and

ε_t_ is the random error term, following a first order autoregressive [AR (1)] process.

It is estimated in STATA Version 15, using the ITSA module^18^, followed by testing for appropriate lag^19^, after adding two sets of extra terms for lockdowns 2 and 3.

### 2.3 SIR Model and related assumptions

In this study we use the SIR model to project the spread of COVID-19 in the absence of lockdowns. The SIR model explains the dynamics of transmission of infectious diseases in a fixed population (of size N). The population is divided into three groups: S (Susceptible), I (Infected), and R (Recovered). Other components of susceptible population are excluded, such as (a) *Exposed*: The model aims to project the epidemic without any intervention; so it is assumed that entire urban population is exposed and will get infected over time; (b) *Quarantined*: In the absence of any contact tracing no quarantine measures will be adopted (c) *Immune*: Herd immunity will increase as population recovers over time. Moreover, there is no vaccine available to assume that certain population can be immune through public health immunization program. Also, the chances of re-infection are not considered to be the major drivers of COVID-19 at the time of writing this article^20,21^.

The model is expressed in terms of three simultaneous non-linear differential equations tracing the time path of the number of susceptible, infected and recovered persons from their starting values (S_0_, I_0_ and R_0_):

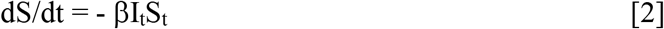

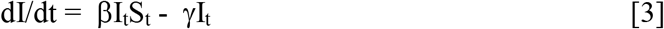

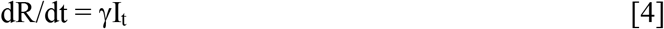

The two parameters of the model, β and γ, represent transmission rate of disease and recovery rate, respectively. The transmission rate is the product of the number of daily contacts of an infected person (c) and the probability that the disease will be transmitted from an infected person to a contact (τ). The recovery rate is the reciprocal of average number of days for recovery.

These parameters define the basic reproduction rate of the disease (R_0_), given by β/γ. If R_0_ is greater than unity, the infection will spread in a self-sustaining manner to become an epidemic; on the other hand, for values of R_0_ lower than unity, the infection will die out in the absence of fresh cases entering the society from outside.

### 2.3 Assumptions of SIR

There are three parameters in the SIR model—daily contacts (c), probability of infection from contact with an infected person (τ), and recovery period (γ). Their combination gives R_0_ (= cτ/γ).

#### 2.3.1 Assumption and survey-based estimates

Although studies on daily contact rates exist for developed countries^22,23^, there is no similar study on daily contacts before lockdown for Indian societies. An online survey conducted by us during the lockdown found mean contact rate *before lockdown* to be 32.21 (95% CI: 23.48, 40.94). Although τ has been estimated to be 2 in China^24,25^, we have taken lower values (τ = 0.75 and 1.00 percent). The value of β (=cτ) is .2416 and 0.3221 (for τ = 0.75 and 1, respectively).

Contact tracing data from 10 early cases in China^26^ showed the mean serial interval (time between successive cases) was 7.5 days with a SD of 3.4 days. A more recent estimate^27^ among 468 cases was 3.96 days with a 4.75 day SD. The outside estimate (mean plus SD) in these studies was 8.71 to 10.9 suggesting a plausible range of 9–11. The WHO-China Joint Mission^28^ reports the median period of recovery to be 14 days, so that γ is 0.0714. Our analysis reveals that, *ceteris paribus*, results are not very sensitive to variations in γ. The value of R_0_ is 3.36 and 4.48, respectively, which appear reasonable for the pre-lockdown period. These projections are referred to as OWNL (for R_0_ = 3.36) and OWNH (for R_0_ = 4.48).

In another set of exercises we have estimated the values of R_0_ based on the Exponential Growth^29^, and Maximum Likelihood^30^ methods, using the R0 package from CRAN^31^. To estimate R_0_, we also require serial interval measures—the time between clinical onset of symptom in primary case and clinical onset of symptom in secondary case^32^. We assume that SI follows a Gamma distribution with mean 7.50 and standard deviation 4.75. A recent study reports the different values of SI for COVID-19^9^. We have taken the highest values^26^, so that our results are likely to err on the higher side. Taking γ to be 0.242921, we have estimated the values of β from the formula R0 = β/γ. The Exponential Growth model yields values of R_0_ and β to be 3.0484 and 0.2177, while the corresponding estimates from the Maximum Likelihood method are 3.0722 and 0.2194. Projections are referred to as EG and ML, respectively.

Till the onset of the fourth lockdown, COVID-19 had been an urban phenomenon—partly because of the restricted testing facilities, and partly because of the containment on migrant workers in urban cities. So we have used the population aged 20 years and above in Class I cities and towns. It has been calculated for 2019 by taking an exponential interpolation of figures from Census 2001 and 2011.

### 2.4 Methodology

The SIR model is run from 6^th^ March to 17^th^ May (the last day of the third lockdown) using alternative values for β and γ. Results generate predicted number of cases in the absence of lockdown. We have four sets of predictions— OWNL, OWNH, EG and ML, corresponding to each model. These give us the counterfactual, and are used to predict the number of COVID cases without lockdown for alternative scenarios. The difference between actual cases and deaths, and estimates using the SIR model (OWNL, OWNH, EG and ML) is an estimate of COVID cases and deaths averted by lockdown; it is used to calculate reduced mortality and pressure on health care system.

The case fatality rate (CFR) is 2.1 percent in Wuhan, but believed to be much lower (0.05) for the rest of the world [33]. Mean of the seven day moving average of fatality rates in India yields a CFR of 0.0298. It is used, along with estimates reported by MoHFW on 21 May 2020^34^ and estimates by Ferguson et al^35^. Parametric values used for all-India estimates are given in Appendix (Table A1). The estimates for required hospital beds, ICU beds, oxygen, and ventilator support are estimated for the peak. State-wise information on health infrastructure is also used in this study^36^.

## 3. Findings and discussion

### 3.1 All India level results

Results of the ITS model is given in Fig. 1 (see also Appendix Table A2). The correlation between predicted and actual values is 0.9869, indicating a very good fit. It may be seen that number of cases tends to increase with time (β_TIME_ = 7.69; Prob: 0.00). The lockdowns seem to have a mixed effect on this trend. The first and second lockdown leads to a level change, with a decline in intercept (Δα = −84.82; prob: 0.00; Δα = −202.90; prob: 0.00); the slope, however, increase sharply (Δβ = 46.47, prob.: 0.00; Δβ = 37.39, prob.: 0.02). After the third lockdown, however, there is an increase in level (Δα = 637.20, prob: 0.00) though slope remains constant (Δβ = 1.64, prob: 0.66).

**Figure 1:**
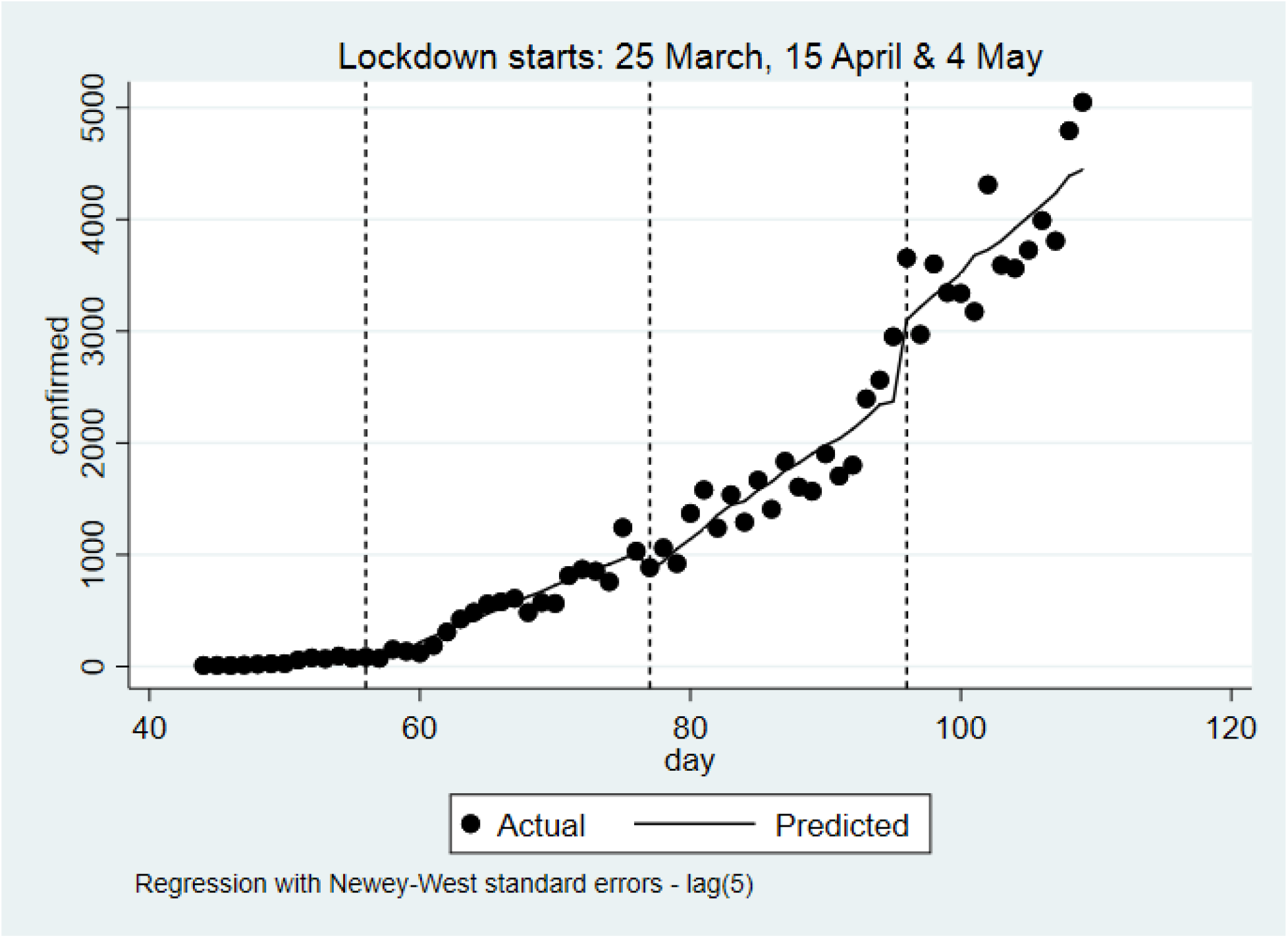
Actual and predicted cases using ITS model.

*Source: Estimated by authors*

Although the lockdowns have failed to prevent the spread of COVID-19, they have succeeded in containing the pace of growth. Results of the SIR model (Figure 2) reveal variations in estimated COVID-19 cases, with our estimates—particularly OWNH—yielding estimates on the higher side. In contrast, results of the SIR models based on estimates from the ML and EG are almost similar.

**Figure 2:**
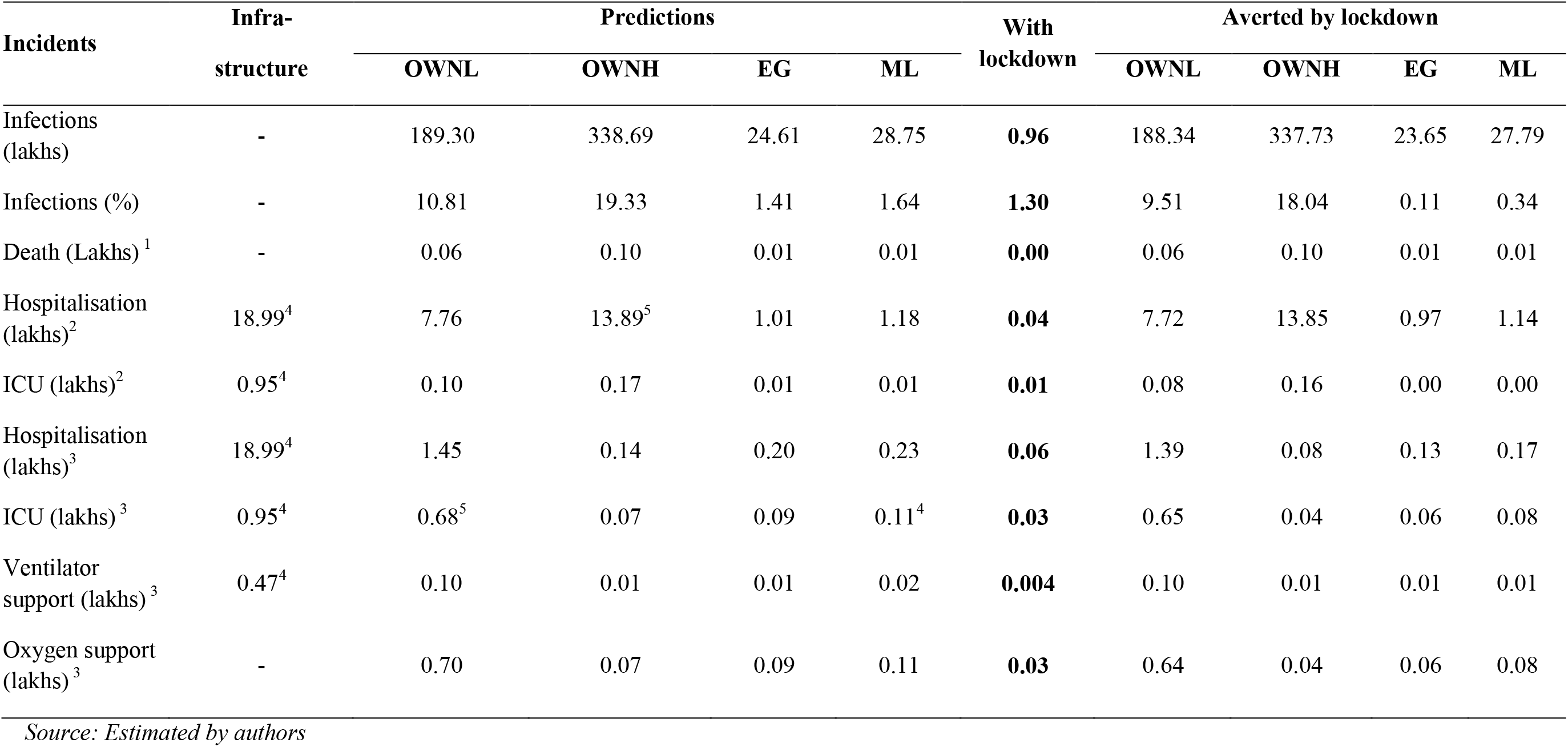
Actual and predicted COVID-19 cases for states.

*Source: Estimated by authors*

The reduction in cases, deaths and pressure on health infrastructure due to lockdowns is summarized in Table 1. While the OWNL and OWNH models indicate a substantial impact of lockdowns, the results from the EG and ML models are quite modest. Another point to note is that the hospital infrastructure does not seem to be strained—only two predictions (OWNL and ML) indicate that there may be a deficit in requirement of ICU beds.

**Table 1:**
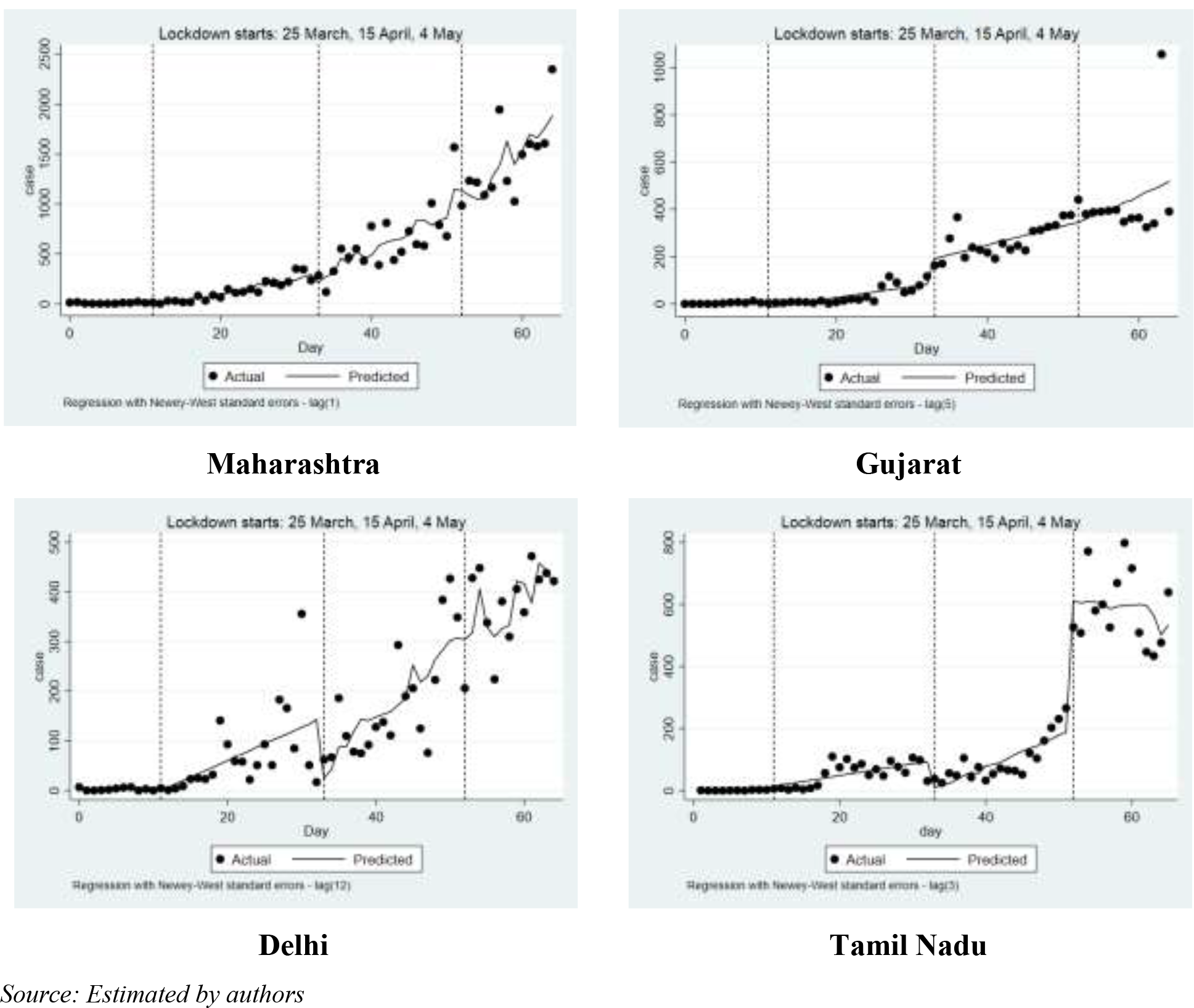
Impact of lockdown on infections, deaths and hospitalization at all-India level.

### 3.2 Analysis for selected states

The national picture hides considerable regional variations. Given the high incidence of cases in some states, it is necessary to also explore the regional picture.

The result of the ITS model (Fig. 2) for Maharashtra indicates failure of the three lockdowns to contain COVID-19; in Gujarat, there is an *increase in level* after the first lockdown, followed by an *increase in slope* after the second lockdown. Only in Delhi and Tamil Nadu, lockdown seems to have achieved limited success—a decrease in level after the Lockdowns 1 and 2, along with a decrease in slope after the third lockdown (Delhi), and a decrease in level after Lockdown 2 (Tamil Nadu).

We have estimated number of COVID-19 cases, mortality, and pressure on hospital infrastructure for these four states assuming that no containment policy had been enforced (Table 2). The estimates have been made based on the SIR model using the parametric assumptions for OWNH and ML models. Results indicate that COVID-19 cases and mortality would have been increased substantially in these states in the absence of policies. A comparison with actual figures, however, indicates that lockdown did not succeed in reducing incidence and mortality substantially (Appendix Table A3). Results also indicate, while hospital beds are sufficient, a major shortage of ICU and ventilator would have occurred in these states.

**Table 2:** Situation in absence of any policies to contain COVID-19—selected states.

| Incidents | OWNH model |  |  |  | Maximum Likelihood (ML) model |  |  |  |
| --- | --- | --- | --- | --- | --- | --- | --- | --- |
|  | Maharashtra | Gujarat | Delhi | Tamil Nadu | Maharashtra | Gujarat | Delhi | Tamil Nadu |
| Infections ('000) | 1,480.18 | 507.28 | 1,345.92 | 487.93 | -32.53 | 7,112.17 | 125.14 | 4,315.66 |
| Infections (% of pop.<br>In Class I cities) | 6.01 | 2.76 | 5.46 | 5.38 | -0.13 | 38.63 | 0.51 | 47.61 |
| Death | 85,663 | 45,761 | 34,050 | 6,809 | -1,167 | 636,898 | 2,310 | 72,253 |
| Hospitalisation cases | 59,103 | 20,302 | 52,659 | 19,504 | 16 | 444,821 | 5,107 | 202,186 |
| ICU cases | 43,246 | 14,855 | 38,531 | 14,271 | 12 | 325,479 | 3,737 | 147,941 |
| Ventilator cases | 6,487 | 2,228 | 5,780 | 2,141 | 2 | 48,822 | 561 | 22,191 |
| Utilisation of hospital<br>bed (%) | 25.50 | 31.30 | 133.46 | 12.55 | 0.01 | 685.80 | 12.94 | 130.13 |
| Utilisation of ICU<br>facilities (%) | 373.23* | 458.08* | 1,989.19* | 183.70* | 0.10 | 10,036.34* | 192.93* | 1,904.25* |
| Utilisation of<br>ventilator support (%) | 111.98* | 137.38* | 586.17* | 55.12 | 0.03 | 3,009.97* | 56.85 | 571.35* |

*Source: Estimated by authors*
**Note:**

### 3.3 Influence of daily testing

The exercise, however, has a major limitation. The SIR model is based upon initial values of the epidemic, and assumes an exponential growth. In India, however, the number of new cases seems to follow a linear fit, with periodic changes in intercept and slope. While this may reflect the impact of containment policies, another possibility is that the trend in cases may reflect, not the spread of disease, but the increase in daily testing. Regression results (Table 3) indicate a positive and statistically relation between testing and new cases. If we eliminate the possible confounding effect of daily testing—by regressing new cases and time upon daily testing, estimating the residuals from each model (thereby eliminating the testing effect), and regressing residual of new cases upon residual of time —a significant and positive relation with time is still observed. It indicates that, even after accounting for the increase in testing, the number of new cases has a statistically significant time trend.

**Table 3:** Relation between new cases, time and testing.

| Variables | Model with actual values |  | Model with residual |  |
| --- | --- | --- | --- | --- |
| | $\beta$ | t | $\beta$ | t |
| Intercept | -1541.01 | -3.27*** | -4.6e<sup>7</sup> | -0.00 |
| TIME | 26.79 | 3.26*** | 26.76 | 3.19*** |
| TESTING | 0.26 | 5.46*** | - | - |
| N |  | 66 | 66 | - |
| F |  | 347.52*** | 10.19*** | - |
| R<sup>2</sup> |  | 0.92 | 0.14 | - |

*Source: Estimated by authors*
**Note:** \*\*\* denotes significance at 1 percent level.

## 4. Conclusion

It is widely accepted that lockdown is a drastic public health measure^37,38^ with far-reaching consequences and adverse health outcomes^39,40^. The adoption of such an extreme step requires careful analysis. Our study indicates that the lockdown has reduced the spread of COVID-19 cases in India. The gains at the national level, however, seem to be modest (except for OWNH model projections); it is also unlikely that the health infrastructure would have been overstrained. Given that COVID-19 in India has varied widely across states, regional analysis is also important. An analysis of the situation in four states with about 68 percent of cases reveals that lockdown may have averted a major health crisis as the health infrastructure (ICU and ventilator support) required for treatment of critically ill patients available in these states would have been inadequate.

In conclusion, we would also like to point out that lockdown is primarily a measure to buy time and create the health infrastructure required to fight COVID-19. This has been done to a limited extent. Consequently, when the lockdown is ultimately lifted, a significant spike can occur at the national level, with the emergence of new hotspots where migrant workers return.

Secondly, estimates—though not conclusions—vary widely depending upon parameters used. The sensitivity to parametric assumptions is a major limitation of existing studies. It calls for more sophisticated calibration of models using parameters relevant for the Indian context.

Finally, our analysis also indicates that the available data may reflect the increase in testing, rather than only the spread of the disease. It implies that the actual number of cases may remain hidden due to limited testing. So, unless we increase daily testing to at least levels comparable with other Asian countries [Singapore (50,368), Bhutan (18,850), Malayasia (14,896), Iran (8,895), Nepal (4,039), Iraq (3,732), Vietnam (2,828), Sri Lanka (2,220), Vietnam (2,828)], estimates of the spread of COVID-19 in India and its analysis may have limitations, reducing the value of policy recommendations and lead to flawed decisions.

## Data Availability

Data on which the research is based is available publicly. Sources have been cited in submission.

https://www.covid19india.org/

## Appendix

**Table A1:** Parametric assumptions used in study for all-India estimates.

| Model | c | $\tau$ | $\beta$ | $\gamma$ | $R_0$ |
| --- | --- | --- | --- | --- | --- |
| OWNL | 32.21 | 0.0075 | 0.2416 | 0.0714 | 3.3821 |
| OWNH | 32.21 | 0.0100 | 0.3221 | 0.0714 | 4.5094 |
| EG model | - | - | 0.2177 | 0.2429 | 3.0484 |
| ML model | - | - | 0.2194 | 0.2429 | 3.0722 |
| Values used (%) | Hospitalization | ICU | Oxygen | Ventilator | CFR |
| Ferguson et al (2020) | 4.1 | 0.88 | - | - | 0.43 |
| Estimated from data | - | - | - | - | 0.0298 |
| MoHFW | 6.39 | 2.94 | 2.94 | 0.45 | - |

*Source: Estimated by authors*

**Table A2:** Regression model using ITS method for India.

| **Variables** | **Newey-West estimates** | **t-ratios** | **Prob. > \|t\|** |
| --- | --- | --- | --- |
| $\alpha$ | -3.24 | -0.16 | 0.87 |
| $\beta_{\text{TESTING}}$ | -0.003 | -0.45 | 0.60 |
| $\beta_{\text{TIME}}$ | 7.69 | 6.04 | 0.00 |
| $\beta_{\text{L1}}$ | 54.17 | 6.55 | 0.00 |
| $\Delta\alpha_{\text{L1}}$ | -84.82 | -2.98 | 0.00 |
| $\Delta\beta_{\text{L1}}$ | 46.47 | 5.11 | 0.00 |
| $\Delta\alpha_{\text{L2}}$ | -202.90 | -1.86 | 0.07 |
| $\beta_{\text{L2}}$ | 91.56 | 4.89 | 0.00 |
| $\Delta\beta_{\text{L2}}$ | 37.39 | 2.49 | 0.02 |
| $\Delta\alpha_{\text{L3}}$ | 637.20 | 3.71 | 0.00 |
| $\beta_{\text{L3}}$ | 105.21 | 3.78 | 0.00 |
| $\Delta\beta_{\text{L3}}$ | 13.64 | 0.44 | 0.66 |
| N | 66 | Lags | 5 |
| F | 440.48 | - | 0.00 |

*Source: Estimated by authors*

**Table A3:** Impact of lockdown in reducing cases, deaths and hospitalization—selected states.

| **OWNH model** | **Maharashtra** | **Gujarat** | **Delhi** | **Tamil Nadu** |
| --- | --- | --- | --- | --- |
| Infections ('000) | 1,480.18 | 507.28 | 1,345.92 | 487.93 |
| Death | 85,663 | 45,761 | 34,050 | 6,809 |
| Hospitalisation | 60,688 | 20,799 | 55,183 | 20,005 |
| **ML model** | **Maharashtra** | **Gujarat** | **Delhi** | **Tamil Nadu** |
| Infections ('000) | - | 7,112.17 | 125.14 | 4,315.66 |
| Death | - | 636,898 | 2,310 | 72,253 |
| Hospitalisation | - | 458,645 | 5,131 | 214,439 |

*Source: Estimated by authors*

## Notes

### Competing Interest Statement

The authors have declared no competing interest.

### Funding Statement

No funding or sponsorship received by us.

### Author Declarations

No patient / clinical data was used. SO IRB approval is not required.

